# The Global Economic Burden of Schizophrenia: An Umbrella Review of Systematic Reviews and Meta-Analyses

**DOI:** 10.1101/2025.06.13.25329410

**Authors:** Attila Imre, Ágota Mészáros, Bertalan Németh, Balázs Nagy, Judit Józwiak-Hagymásy, Giacomo Cecere, Philipp Homan, Iris E.C. Sommer, Balázs Babarczy

## Abstract

**Background:** Existing studies on the economic impact of schizophrenia are fragmented and vary in methodology, hindering an ample understanding of the total economic burden.

**Aims:** To quantify the global economic burden of schizophrenia by synthesizing findings from systematic reviews and meta-analyses, and to assess variations by cost category, world region and income level.

**Method:** We conducted a protocol-driven umbrella review registered on PROSPERO (CRD42024504092). A systematic search of MEDLINE, EMBASE, Cochrane Library, and APA PsycINFO identified reviews reporting annual per-patient monetary cost estimates for schizophrenia. Monetary values were adjusted for inflation and converted to 2024 US dollars.

**Results:** Twenty-six systematic reviews involving 152 primary studies were included. The median annual per-patient total cost of schizophrenia was $33,236 (mean $47,872), with direct costs accounting for $23,126 (medical: $19,543; ancillary: $1,152) and indirect costs for $21,333. Costs were varied by income level: in high-income countries, the median total cost was $34,175, compared to $3,345 in upper-middle-income and $3,452 in lower-middle-income countries. Regional disparities were substantial, with Europe & Central Asia reporting the highest median costs among high-income settings. Data from low- and middle-income regions were limited or absent. Methodological heterogeneity, lack of standardization in cost reporting, and underrepresentation of low-resource settings limit generalizability.

**Conclusions:** Schizophrenia poses a significant and unequally distributed global economic burden, driven by hospitalization, long-term care, and productivity losses. To support equitable policy and resource allocation, future research must adopt standardized costing frameworks and expand data collection into low-resource and understudied regions.

## Introduction

Schizophrenia is a chronic and often severe psychiatric disorder that affects approximately 20 million people worldwide, profoundly disrupting perception, thought, affect and behaviour.^1,2^ Its clinical hallmarks - hallucinations, delusions, disorganized thinking and speech - are frequently accompanied by cognitive deficits and impaired social functioning, placing enormous strain on patients, families, and communities alike.^3,4^ In addition to its clinical burden, schizophrenia is associated with substantial long-term disability, markedly reduced quality of life, and a life expectancy shortened by 10-20 years compared with the general population.^5-7^ These adverse outcomes translate into a considerable global burden: recent estimates attribute up to 1.3 percent of total disability-adjusted life years lost to schizophrenia and related psychoses, underscoring its public health importance.^8,9^

The economic consequences of schizophrenia are multifaceted. Direct healthcare costs encompass inpatient and outpatient services, long-term residential care, emergency visits, and pharmacotherapy, with antipsychotic medications and hospital stays constituting the largest single expenditures in many settings.^10,11^ Beyond these medical expenses there are indirect costs: lost productivity from unemployment, absenteeism and presenteeism; premature mortality; and the unpaid labour of family members and informal caregivers often equal to or greater than direct costs.^12,13^ Families frequently shoulder a disproportionate share of the financial burden, experiencing reduced income, increased out-of-pocket expenses, and substantial psychosocial stress^12^. Moreover, stigma and discrimination not only worsen clinical outcomes but also erect barriers to accessing care and social supports, further amplifying the disorder’s economic impact.^14^

Although a growing number of systematic reviews and meta-analyses have examined specific cost components or evaluated the economic implications of individual interventions like long-acting injectable antipsychotics, these syntheses remain fragmented. Differences in study design, costing methodologies, choice of cost categories, currency adjustments, and time horizons hinder meaningful comparisons across regions and over time. As a result, policymakers and healthcare planners lack a comprehensive, harmonized understanding of schizophrenia’s total economic footprint.

Umbrella reviews, which systematically collate and assess evidence from multiple systematic reviews and meta-analyses, offer a powerful means to bridge this gap by providing a high-level overview of the existing literature, identifying areas of concordance and discordance, and pinpointing critical evidence gaps^15^. By integrating findings across heterogeneous studies, umbrella reviews can elucidate the full scope of economic burden, reveal under-studied cost drivers, and guide both research priorities and resource allocation strategies^16^. To date, however, no umbrella review has synthesized the global economic burden of schizophrenia across diverse healthcare and socioeconomic settings.

In this context, we conducted an umbrella review to map and quantify the costs associated with schizophrenia, encompassing both direct and indirect components, across high-, middle-, and low-income countries. Our objectives were to (1) summarize the per-patient per-year (PPPY) costs; (2) examine variations by World Bank country income group and geographic region; (3) deconstruct cost estimates into meaningful categories. By offering the first overview of schizophrenia’s economic impact across settings, we aim to inform clinicians, health economists, and policymakers in their efforts to allocate resources effectively, design cost-effective interventions, and ultimately mitigate the substantial societal burden of this debilitating disorder.^17,18^

## Methods

For this umbrella review, a systematic review protocol was developed and registered on PROSPERO (CRD42024504092).^19^ The review was conducted in accordance with the PRISMA guidelines.^20^

Studies were selected based on predefined eligibility criteria using the PICOS framework.^21^ Studies were eligible if they were systematic reviews or meta-analyses reporting on the economic burden of schizophrenia or schizoaffective disorder, including direct and/or indirect costs. Reviews were included if they provided a reproducible search strategy and adhered to systematic review methods. Only studies published in English were considered. No geographic restrictions were applied. Reviews without a clear description of search and selection methods were excluded.

Two literature searches were conducted to identify reviews examining the economic burden of schizophrenia. The electronic databases searched included MEDLINE (via PubMed), EMBASE, the Cochrane Library of Systematic Reviews, and APA PsycINFO.

The first search for MEDLINE, EMBASE and the Cochrane Library was conducted on December 4^th^, 2023, and APA PsycINFO on December 11^th^, 2023. A second search was performed on April 9^th^, 2025, with no date restriction. The search strategy was constructed using a combination of concepts related to schizophrenia, economic burden, and study type. A systematic approach was undertaken to develop the search queries. Relevant keywords for each concept were identified by the review authors and supplemented with terms from the Medical Subject Headings (MeSH) and the Unified Medical Language System (UMLS) metathesaurus. The search terms included synonyms and related terms for schizophrenia, economic burden, cost of illness, systematic review, and meta-analysis. The complete search queries for the targeted databases are provided in the Appendix.

After removing duplicates, titles and abstracts from the first search were screened independently by two reviewers using Covidence^22^. Titles and abstracts from the second search were screened by AI. Full texts were screened in the same manner. All papers were evaluated as published, including supplementary materials and online appendices when available. The authors were not contacted for additional information. Disagreements were resolved through discussion or consultation with a senior reviewer with domain expertise (BB, BN).

Data were extracted using Microsoft Excel® and included review-level characteristics (e.g., author, year, objective, databases searched, population, interventions, outcomes). However, as many reviews lacked consistent or detailed cost breakdowns, we deviated from the original protocol by systematically extracting structured cost data directly from peer-reviewed primary studies cited within the included reviews. Only studies reporting annual per-patient monetary cost values were included in this extraction. Ratios or differences between two different groups of patients (e.g., two different treatment groups) were not extracted.

The methodological quality of included reviews and meta-analyses was assessed by an individual reviewer using the AMSTAR 2 checklist.^23^ The risk of bias assessment was double-checked by a second reviewer. Confidence ratings were calculated according to Shea et al.^23^

Following method two described by Turner et al. ^24^, we adjusted extracted monetary values to 2024 US dollars. First, we inflated local currencies to the year 2024, using the Consumer Price Index^25^ data provided by the World Bank for non-European countries and the Harmonised Index of Consumer Prices (HICP)^26^ published by the Eurostat. For data extracted for Taiwan, we used the consumer price index published by National Statistics, Republic of China.^27^ Second, we exchanged local currencies to 2024 US dollars we utilized exchange rates^28^ published by the World Bank.

The overlap between primary studies included in the reviews was characterized by corrected covered area (CCA) as described by Pieper et al.^29^ For this we built a citation matrix in which primary publications are listed in rows and the reviews included in the umbrella review are listed in columns. Based on the citation matrix, we calculated how many times a given study is cited in the included reviews. The equation for CCA is as follows:

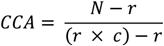

where *N* is the number of study occurrences across all reviews (inclusive of duplicates), *r* is the number of unique primary studies, and *c* is the number of reviews.

Given the substantial heterogeneity in cost definitions, population characteristics, and outcome reporting across the included reviews, a meta-analytic synthesis was not feasible. Therefore, we employed a structured narrative synthesis to summarize cost estimates across settings and categories. Results were tabulated and described by cost type, income group, and geographic region. To explore variability in economic burden estimates, we conducted stratified analyses based on World Bank income group and region. These groupings allowed for descriptive comparisons of cost patterns across settings. As a sensitivity analysis, we repeated the primary cost summaries while restricting to studies published after 2000 to evaluate the robustness of estimates over time.

We did not formally assess the risk of reporting bias in the primary studies, due to the secondary nature of this umbrella review and the lack of consistent reporting bias evaluations in the included reviews. Certainty of the evidence was not assessed also because this umbrella review focused on economic burden estimates, which lacked standardized outcome measures across reviews.

## Results

A total of 1,647 records were identified through multiple sources: MEDLINE (n = 311), EMBASE (n = 586), Cochrane (n = 520), APA PsycINFO (n = 229), and hand search (n = 1). Out of 1,126 hits from MEDLINE, EMBASE, and APA PsycINFO (databases that index articles in multiple languages) 1,190 records were in English (94.62%). After removing 200 duplicates, 1,447 records underwent title and abstract screening. Of these, 1,276 were excluded, leaving 171 reports for full-text retrieval. One report could not be retrieved, resulting in 170 reports assessed for eligibility. A total of 144 reports were excluded: 11 were not focused on schizophrenia, 77 did not report monetary economic burden, and 56 were not systematic reviews. Ultimately, 26 systematic reviews and 152 unique primary studies were included in the final analysis. The study selection process is illustrated in the PRISMA flowchart (Figure 1). No statistical synthesis or meta-analysis was conducted. As such, effect estimates and measures of precision are not reported. All findings are presented descriptively based on extracted monetary cost data.

**Figure 1.**
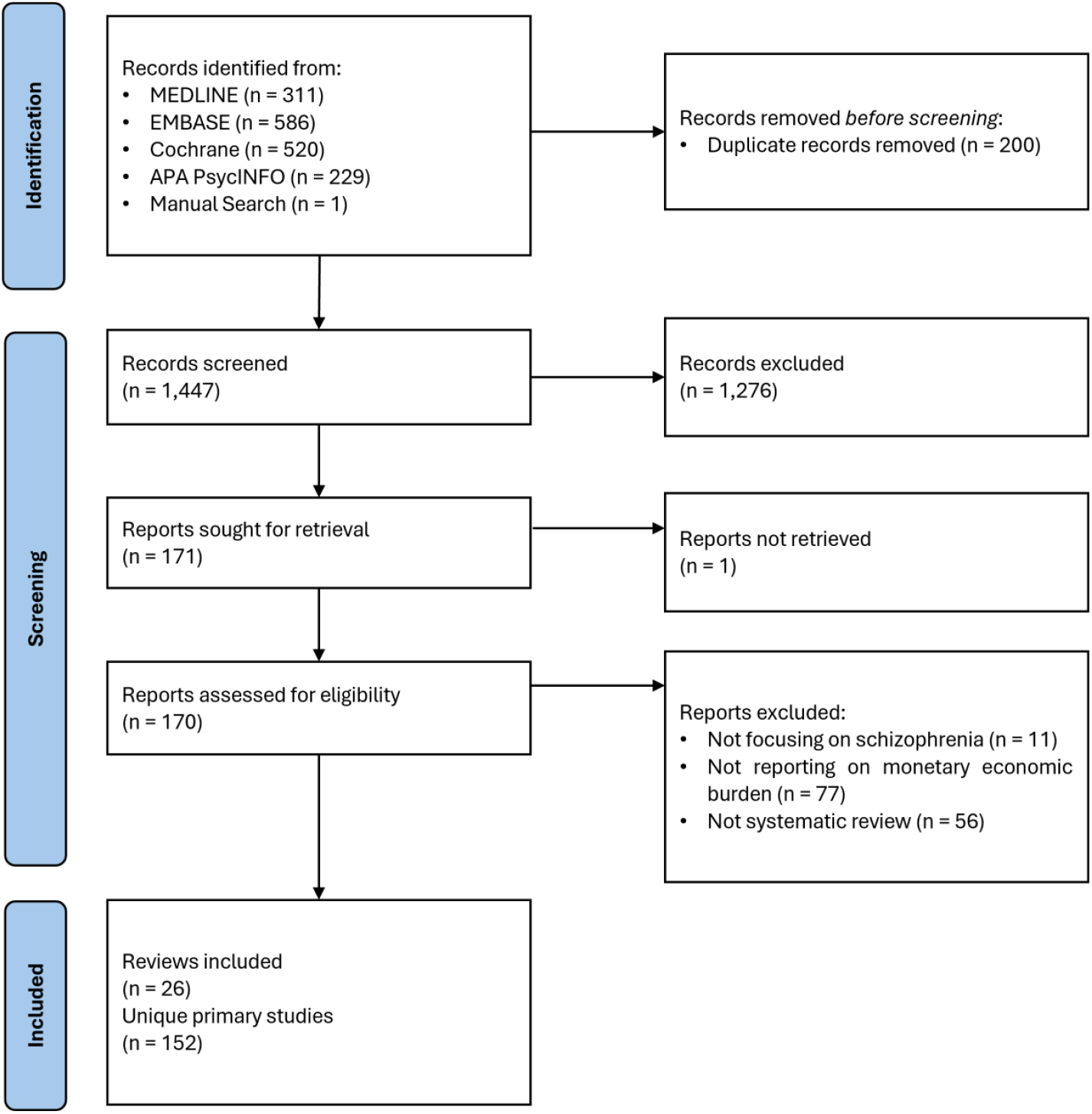
PRISMA flow diagram of study identification, screening, eligibility and inclusion in the umbrella review.

Table 1 summarizes the key characteristics of the 26 included reviews. Seventeen (65%) were systematic literature reviews (SLRs)^13,30-45^, 6 (23%) were SLRs with meta-analyses (SLR & MA)^46-51^, 2 (8%) were targeted literature reviews (TLRs)^52,53^, and 1 (4%) was an SLR with an embedded cost-of-illness study (SLR & COI)^54^. Seventeen reviews (65%) had a global scope^13,30-34,39,41-44,46-51^; others focused on the United States (n = 4)^37,40,45,52^, Europe (n = 3)^35,36,38^, Italy (n = 1)^54^, or a group of 10 countries (n = 1)^53^. Commonly searched databases included MEDLINE (n = 24), EMBASE (n = 17), APA PsycINFO (n = 15), CINAHL (n = 5), Cochrane Library (n = 5), and Scopus (n = 4).

**Table 1.**
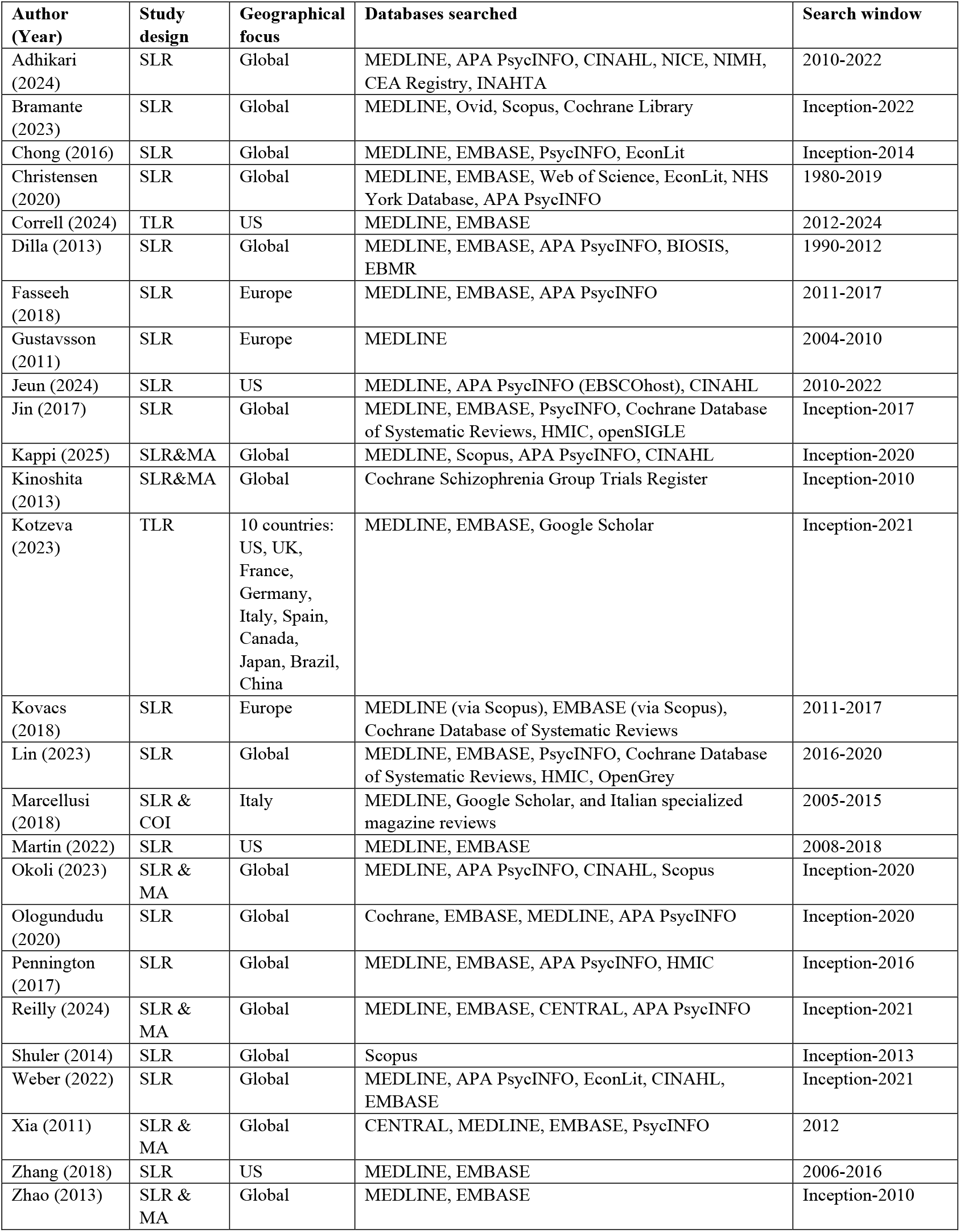
Characteristics of included reviews.

Figure 2 displays the search coverage periods for the included reviews. The earliest searches ended in 2010 and were conducted by Zhao et al. (2013)^46^, Gustavsson et al. (2011)^36^, and Kinoshita et al. (2013)^48^. The review with the latest search was by Correll et al. (2024)^52^, which concluded in 2024. Of the included reviews, 14 searched from database inception^13,31,32,41-44,46-51,53^, while 12 specified a defined start year^30,33-40,45,52,54^. Among the reviews with a defined start year, the mean search span was 12.42 years (SD = 9.59). On average, 1.73 years (SD = 1.37) elapsed between the end of the search period and the publication year.

**Figure 2.**
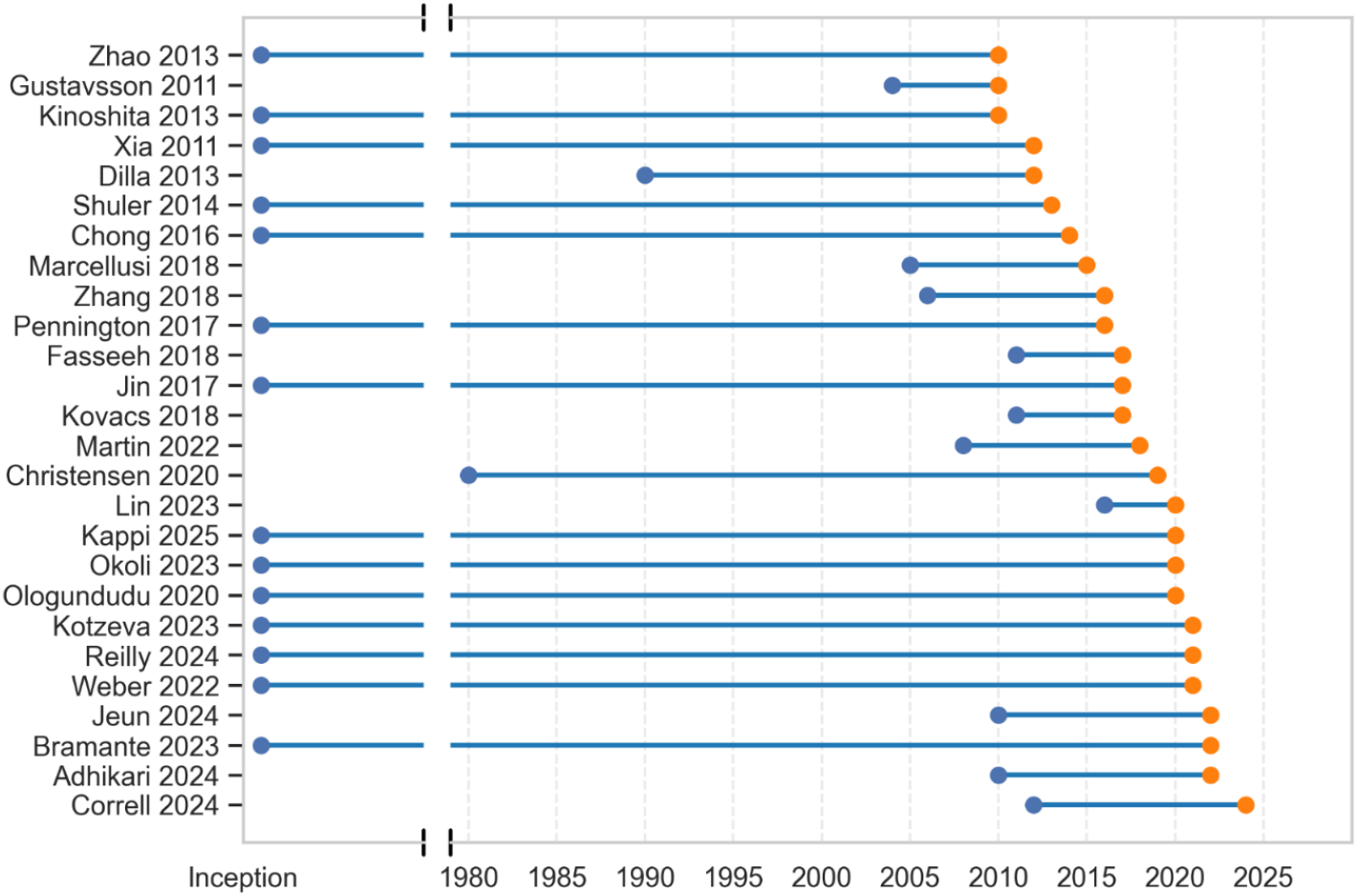
Search coverage periods of included reviews. Each horizontal line represents a review, with the start and end points indicating the range of years covered by its search strategy. Orange dots mark the final year of coverage. Reviews are ordered by their search end year (listed alongside author names and publication year).

The calculated corrected covered area (CCA) was 3.32%, which comprised 278 study occurrences (N) across 26 reviews (c), with 152 unique primary studies (r). This falls within the “slight overlap” category (0%-5%) as defined by Pieper et al.^29^

### Quality assessment

The methodological quality of the 26 included systematic reviews was assessed using the AMSTAR 2 checklist.^23^ Of these, 22 reviews (85%) were rated as having “critically low” confidence, and four reviews (15%) as “low” confidence. Several AMSTAR 2 items were frequently unmet. A comprehensive literature search strategy (Q4) was completely satisfied in only 1 review (3.8%), justification of excluded studies (Q7) in 3 reviews (11.5%), and appropriate risk of bias assessment for included studies (Q9) in 3 reviews (11.5%). Protocol registration (Q2) and adequate description of included studies (Q8) were each satisfied in 4 reviews (15.4%). Duplicate study selection (Q5) and data extraction (Q6) were satisfied in 17 (65.4%) and 11 (42.3%) reviews, respectively.

Among the six reviews that included meta-analyses, five (83.3%) used appropriate statistical synthesis methods (Q11), four (66.7%) assessed publication bias (Q15), and three (50%) evaluated the influence of risk of bias on meta-analytic results (Q12). Reporting of potential conflicts of interest or funding sources (Q16) was the most consistently satisfied item, reported in 24 reviews (92.3%). Detailed results are provided in Supplementary Table 6.

### High level cost breakdown

As shown in Figure 3, annual per-patient costs (2024 US$ PPPY) varied across high-level categories. Total cost (reported as such in the primary studies, n = 43) had a median of $33,236 (IQR = $14,737-$52,919) and a mean of $47,872 (SD = $63,166). Overall direct cost (n = 276) was reported with a median of $23,126 (IQR $10,929-$36,584) and a mean of $30,689 (SD $31,540); within this, direct medical cost (n = 68) showed a median of $19,543 (IQR $9,883-$31,123) and a mean of $25,272 (SD $23,642), while direct ancillary cost (n = 19) was substantially lower (median $1,152, IQR $226-$5,996; mean $4,189, SD $5,495). Overall indirect cost (n = 27) had a median of $21,333 (IQR $8,994-$32,323) and a mean of $24,159 (SD $20,325).

**Figure 3.**
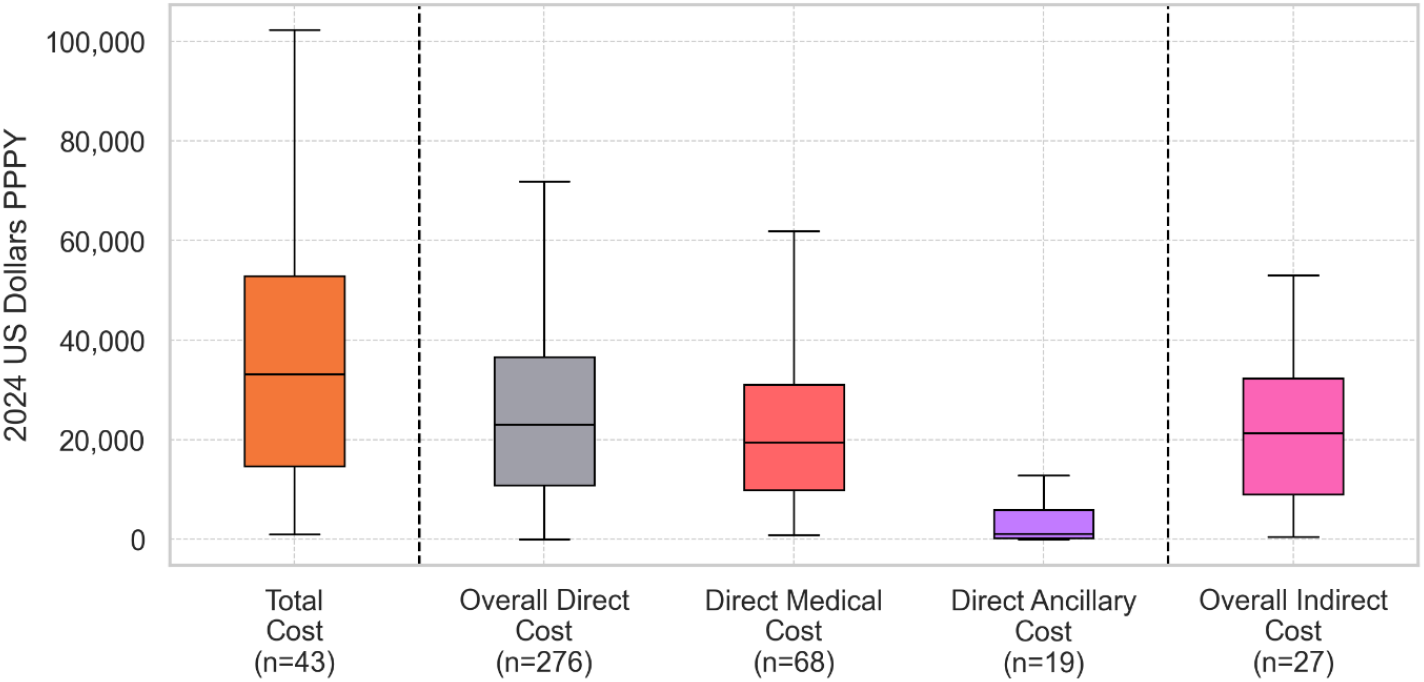
Distribution of annual costs per person by cost category (2024 USD PPPY). Boxplots show the median, interquartile range for total costs, overall direct costs, direct medical costs, direct ancillary costs and overall indirect Costs. Sample sizes (n) reflect the number of cost estimates available in each category as reported in primary studies. PPPY: Per person per year

Costs stratified by World Bank income group and region are presented in Table 2. In high-income countries (n = 36 total-cost estimates), the median total cost was $34,175 (IQR $26,525-$62,770; mean $56,353, SD $65,819), the median overall direct cost $23,985 (IQR $12,082-$36,822; mean $32,173, SD $31,688; n = 262), and the median overall indirect cost $24,436 (IQR $19,353-$37,360; mean $29,072, SD $19,344; n = 22). Within this group, North America (n = 10) showed a median total cost of $33,692 (IQR $29,484-$34,474; mean $35,458, SD $15,499), Europe & Central Asia (n = 19) a median of $47,801 (IQR $20,415-$79,001; mean $71,597, SD $86,723), and East Asia & Pacific (n = 7) a median of $38,361 (IQR $30,242-$45,266; mean $44,826, SD $27,016).

**Table 2.**
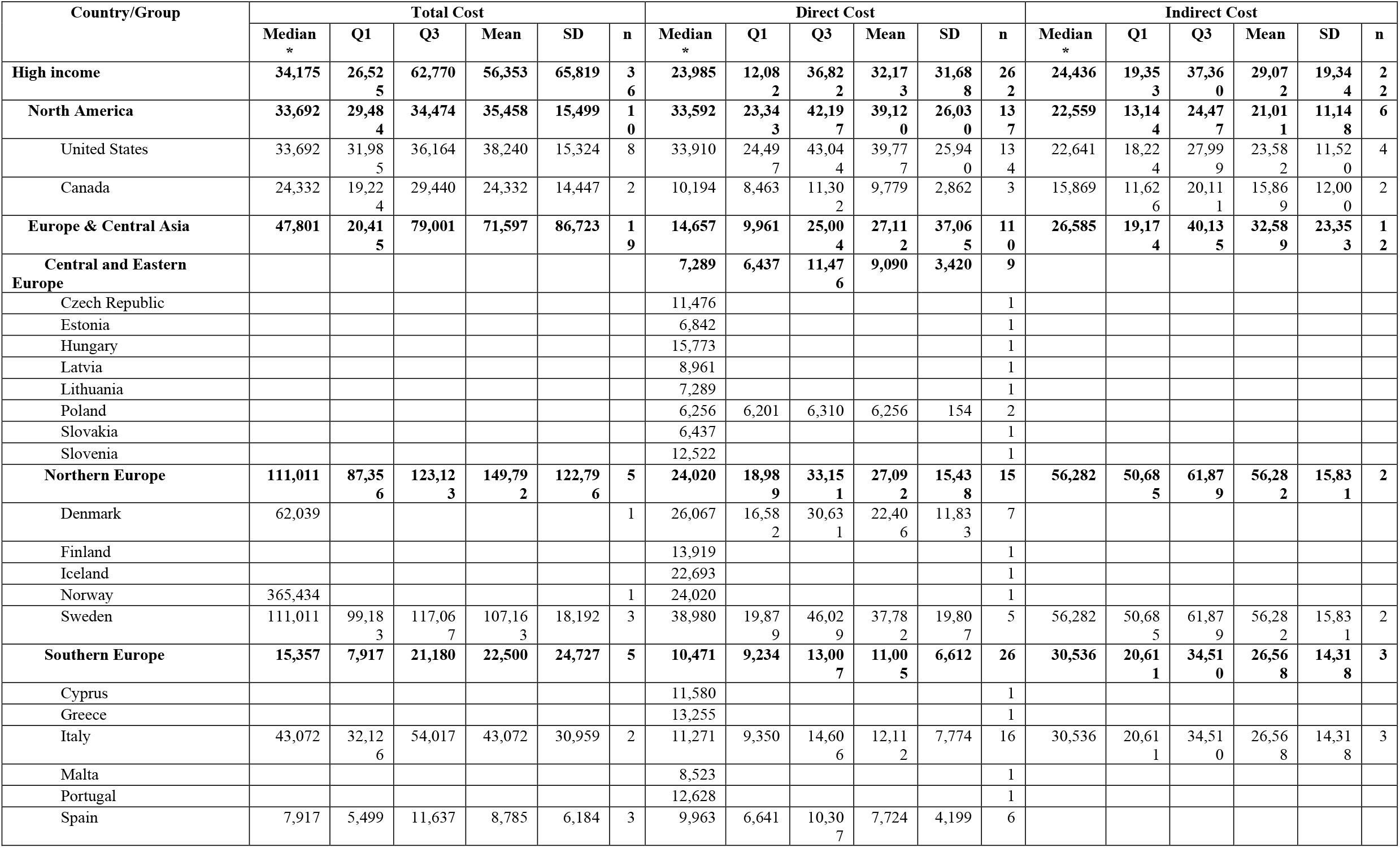
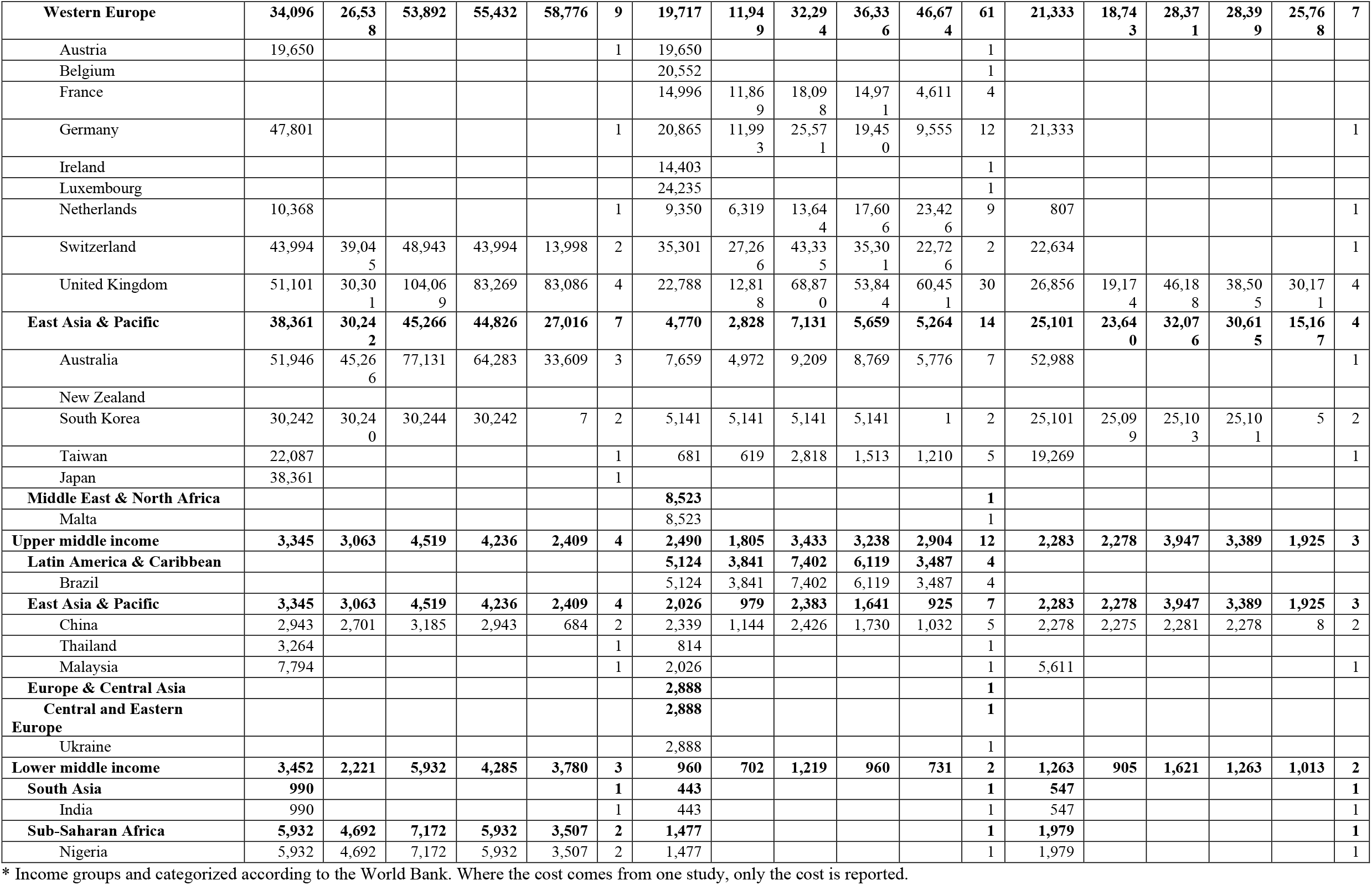
Total costs, direct costs and indirect costs per year per country, region and income group. Costs are expressed as 2024 US dollars.

Upper-middle-income countries (n = 4) had substantially lower costs: median total cost $3,345 (IQR $3,063-$4,519; mean $4,236, SD $2,409), median overall direct cost $2,490 (IQR $1,805-$3,433; mean $3,238, SD $2,904; n = 12), and median overall indirect cost $2,283 (IQR $2,278-$3,947; mean $3,389, SD $1,925; n = 3). Lower-middle-income countries (n = 3) reported a median total cost of $3,452 (IQR $2,221-$5,932; mean $4,285, SD $3,780); direct costs were low (median $960, IQR $702-$1,219; mean $960, SD $731; n = 2) and indirect costs modest (median $1,263, IQR $905-$1,621; mean $1,263, SD $1,013; n = 2). In Sub-Saharan Africa (n = 2), both estimates came from Nigeria, yielding a median total cost of $5,932 (IQR $4,692-$7,172; mean $5,932, SD $3,507); direct and indirect costs were minimal (medians $1,477 and $1,979, respectively; n = 1 each).

### Direct medical costs

Costs for direct medical services as reported in primary studies are detailed in Figure 4. Cost categories are reported as in the primary studies extracted. Among inpatient care, unspecified hospitalization (n = 79) had a median cost of $6,465 (IQR $2,585-$15,310) and a mean of $12,152 (SD $14,377). Long-term care (n = 39) followed with a median of $4,981 (IQR $1,078-$11,002) and a mean of $12,003 (SD $30,819), while general inpatient care (n = 179) was reported with a median of $4,398 (IQR $1,145-$13,989) and a mean of $12,818 (SD $25,040). Psychiatric hospitalization (n = 48) carried a median cost of $2,798 (IQR $1,782-$4,352) and a mean of $14,389 (SD $59,104), whereas non-psychiatric hospitalization (n = 5) was reported as substantially lower (median $1,388; IQR $904-$1,561; mean $1,151; SD $549) and acute hospitalization (n = 29) lower still (median $905; IQR $759-$1,024; mean $941; SD $425).

**Figure 4.**
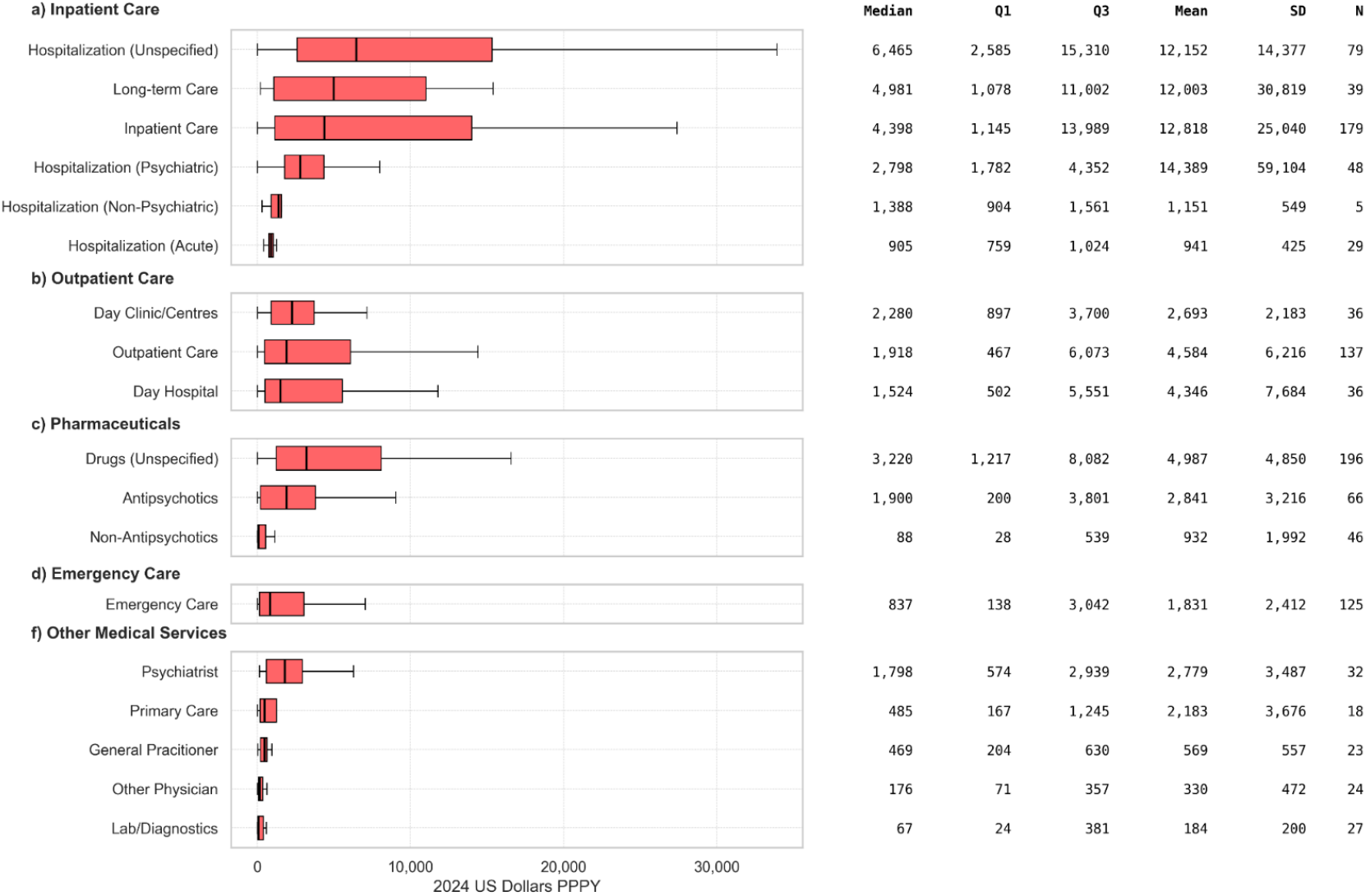
Annual per-person costs by healthcare service category (2024 USD PPPY). Boxplots show the distribution of costs across subcategories within (a) Inpatient Care, (b) Outpatient Care, (c) Pharmaceuticals, (d) Emergency Care, and (f) Other Medical Services. SD: Standard Deviation; N: Number of Cost Estimates

Outpatient services also spanned a broad range: day clinics/centres (n = 36) had a median of $2,280 (IQR $897-$3,700) and a mean of $2,693 (SD $2,183), outpatient visits (n = 137) a median of $1,918 (IQR $467-$6,073) and a mean of $4,584 (SD $6,216), and day hospital services (n = 36) a median of $1,524 (IQR $502-$5,551) with a mean of $4,346 (SD $7,684).

Unspecified drug costs (n = 196) had a median of $3,220 (IQR $1,217-$8,082) and a mean of $4,987 (SD $4,850); antipsychotics (n = 66) a median of $1,900 (IQR $200-$3,801) and a mean of $2,841 (SD $3,216); and non-antipsychotics (n = 46) a median of $88 (IQR $28-$539) and a mean of $932 (SD $1,992).

Emergency care (n = 125) had a median cost of $837 (IQR $138-$3,042) and a mean of $1,831 (SD $2,412). Other medical services were reported lower overall: psychiatrist services (n = 32) showed a median of $1,798 (IQR $574-$2,939) and a mean of $2,779 (SD $3,487), primary care (n = 18) a median of $485 (IQR $167-$1,245) and a mean of $2,183 (SD $3,676), general practitioner visits (n = 23) a median of $469 (IQR $204-$630) and a mean of $569 (SD $557), other physician services (n = 24) a median of $176 (IQR $71-$357) and a mean of $330 (SD $472), and laboratory/diagnostic procedures (n = 27) a median of $67 (IQR $24-$381) with a mean of $184 (SD $200).

### Direct ancillary costs

Direct ancillary costs as reported in primary studies are presented in Figure 5. Case management (n = 4) had a median of $1,263 (IQR $788-$2,151) and a mean of $1,676 (SD $1,271), and rehabilitation services (n = 40) had a median of $353 (IQR $77-$2,570) and a mean of $3,060 (SD $5,933).

**Figure 5.**
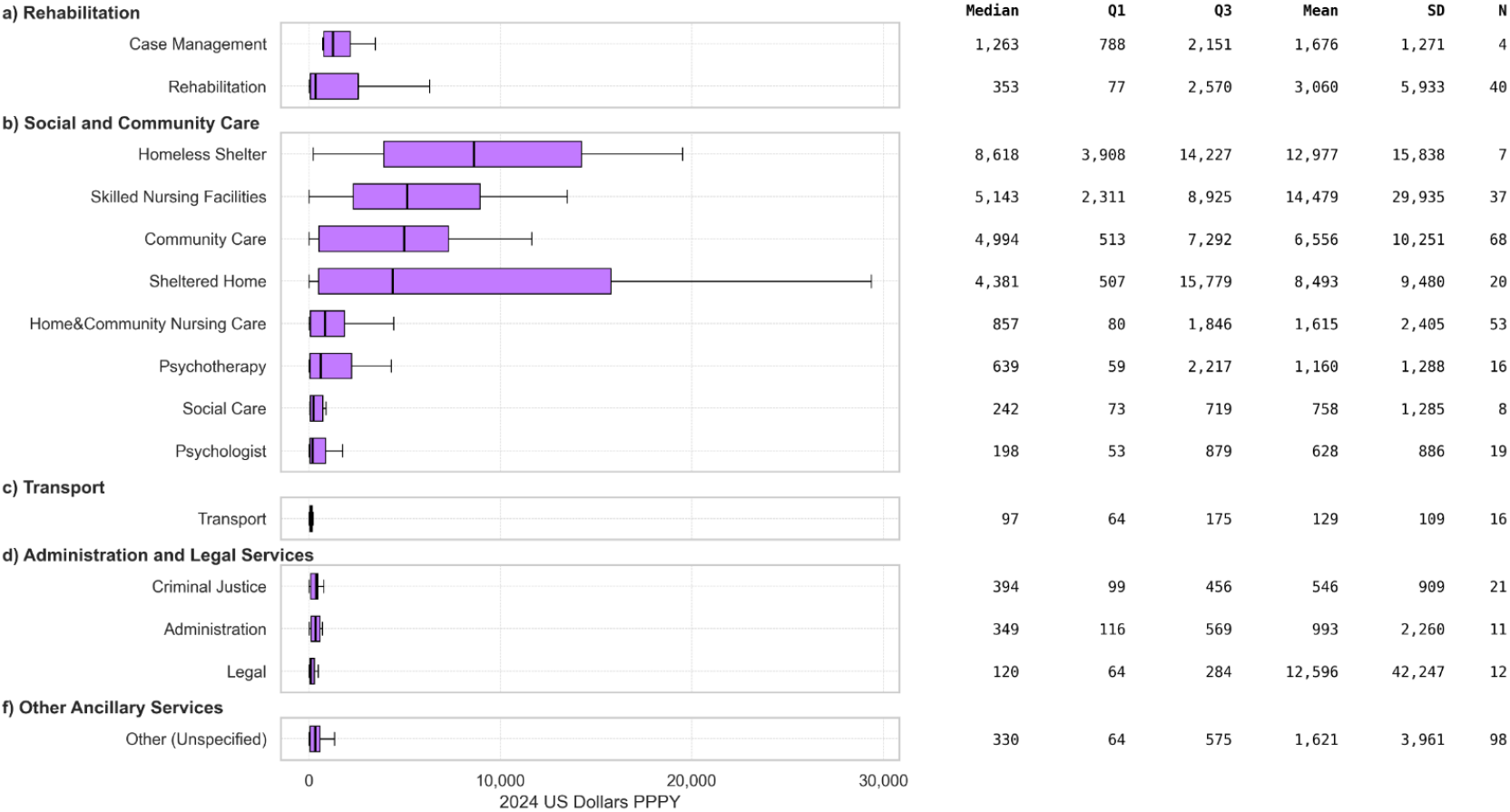
Annual per-person non-medical costs by category (2024 USD PPPY). Boxplots show cost distributions across subcategories of (a) Rehabilitation, (b) Social and Community Care, (c) Transport, (d) Administration and Legal Services, and (f) Other Ancillary Services. SD: Standard Deviation; N: Number of Cost Estimates

Homeless shelter services (n = 7) were reported with a median cost of $8,618 (IQR $3,908-$14,227) and a mean of $12,977 (SD $15,838); skilled nursing facilities (n = 37) a median of $5,143 (IQR $2,311-$8,925) and a mean of $14,479 (SD $29,935); community care (n = 68) a median of $4,994 (IQR $513-$7,292) and a mean of $6,556 (SD $10,251); sheltered homes (n = 20) a median of $4,381 (IQR $507-$15,779) and a mean of $8,493 (SD $9,480); home & community nursing care (n = 53) a median of $857 (IQR $80-$1,846) and a mean of $1,615 (SD $2,405); psychotherapy (n = 16) a median of $639 (IQR $59-$2,217) and a mean of $1,160 (SD $1,288); social care (n = 8) a median of $242 (IQR $73-$719) and a mean of $758 (SD $1,285); and psychologist services (n = 19) a median of $198 (IQR $53-$879) and a mean of $628 (SD $886).

Transport (n = 16) was reported with a median cost of $97 (IQR $64-$175) and a mean of $129 (SD $109). Criminal justice services (n = 21) had a median of $394 (IQR $99-$456) and a mean of $546 (SD $909); administrative services (n = 11) a median of $349 (IQR $116-$569) and a mean of $993 (SD $2,260); and legal services (n = 12) a median of $120 (IQR $64-$284) and a mean of $12,596 (SD $42,247). Other unspecified ancillary services (n = 98) had a median of $330 (IQR $64-$575) and a mean of $1,621 (SD $3,961).

### Indirect costs

Indirect costs as reported in primary studies are detailed in Figure 6. Morbidity-related cost (n = 9) had a median of $17,408 (IQR $9,351-$24,353) and a mean of $16,260 (SD $10,399). Productivity loss (n = 42) had a median of $8,091 (IQR $1,322-$27,135) and a mean of $17,226 (SD $20,932). Informal care cost (n = 10) had a median of $3,638 (IQR $1,909-$14,557) and a mean of $7,739 (SD $7,327). Mortality-related cost (n = 11) had a median of $1,306 (IQR $647-$2,372) and a mean of $99,532 (SD $325,854). Absenteeism (n = 8) had a median of $1,304 (IQR $432-$3,606) and a mean of $5,180 (SD $10,466). Presenteeism (n = 6) had a median of $306 (IQR $169-$1,623) and a mean of $1,156 (SD $1,630).

**Figure 6.**
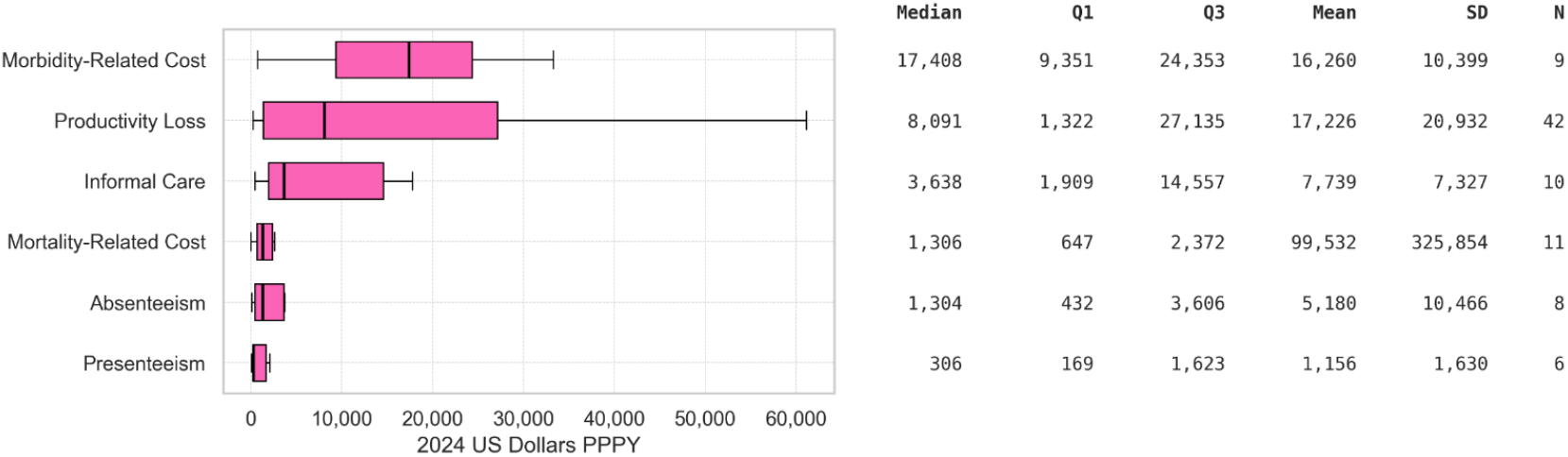
Annual per-person indirect costs by category (2024 USD PPPY). Boxplots show distributions for Morbidity-Related Costs, Informal Care, Productivity Loss, Mortality-Related Costs, Absenteeism, and Presenteeism. SD: Standard Deviation; N: Number of Cost Estimates

### Comparative analysis of costs estimates in studies published after 2000

When restricted to reports published after 2000 (Supplementary Table S7), cost estimates remained broadly similar with modest shifts. In high-income countries (n = 26 total-cost estimates), the median annual total cost was $34,175 (IQR $30,240–$71,333; mean $64,604, SD $75,026), compared with $34,175 (IQR $26,525–$62,770; mean $56,353, SD $65,819; n = 36) for the full dataset. The median overall direct cost increased to $25,697 (IQR $13,713–$36,886; mean $32,714, SD $30,959; n = 206) versus $23,985 (IQR $12,082–$36,822; mean $32,173, SD $31,688; n = 262), and the median overall indirect cost rose to $24,807 (IQR $21,658–$43,427; mean $34,117, SD $20,774; n = 14) versus $24,436 (IQR $19,353–$37,360; mean $29,072, SD $19,344; n = 22). In upper-middle-income countries there was no change in estimates.

Among lower-middle-income countries (n = 2), the median total cost decreased to $2,221 (IQR $1,605–$2,837; mean $2,221, SD $1,741) from $3,452 (IQR $2,221–$5,932; mean $4,285, SD $3,780; n = 3), while median overall direct ($960; IQR $702–$1,219; mean $960, SD $731; n = 2) and indirect costs ($1,263; IQR $905–$1,621; mean $1,263, SD $1,013; n = 2) remained unchanged.

### Regional differences

When compared by income group (Table 2), median annual total costs rose steeply with country income. In high-income countries (n = 36), the median total cost was $34,175 (IQR $26,525-$62,770; mean $56,353, SD $65,819), versus $3,345 (IQR $3,063-$4,519; mean $4,236, SD $2,409; n = 4) in upper-middle-income countries and $3,452 (IQR $2,221-$5,932; mean $4,285, SD $3,780; n = 3) in lower-middle-income countries. Median overall direct costs followed the same pattern ($23,985; IQR $12,082-$36,822; mean $32,173 in high income, $2,490; IQR $1,805-$3,433; mean $3,238 in upper-middle, and $960; IQR $702-$1,219; mean $960 in lower-middle), as did median indirect costs ($24,436; IQR $19,353-$37,360; mean $29,072; $2,283; IQR $2,278-$3,947; mean $3,389; and $1,263; IQR $905-$1,621; mean $1,263, respectively).

Within high-income settings, the median total cost reported from North America (n = 10) was $33,692 (IQR $29,484-$34,474; mean $35,458, SD $15,499), in East Asia & Pacific (n = 7) $38,361 (IQR $30,242-$45,266; mean $44,826, SD $27,016), and in Europe & Central Asia (n = 19) $47,801 (IQR $20,415-$79,001; mean $71,597, SD $86,723). Subregional medians within Europe & Central Asia were $111,011 in Northern Europe (n = 5; IQR $87,356-$123,123; mean $149,792, SD $122,796), $15,357 in Southern Europe (n = 5; IQR $7,917-$21,180; mean $22,500, SD $24,727) and $34,096 in Western Europe (n = 9; IQR $26,538-$53,892; mean $55,432, SD $58,776). No total-cost estimates were identified for Middle East & North Africa

### Cost drivers by component

At the median level, the reported level of direct medical costs was equal to 59% of the value reported for total cost (median $19,543 of $33,236), direct ancillary costs were equal to 3% (median $1,152), and indirect costs were equal to 64% (median $21,333). At the mean level, direct medical amounted to 53% of the value of total cost (mean $25,272 of $47,872), direct ancillary costs to 9% (mean $4,189), and indirect costs to 51% (mean $24,159).

The five cost components with the highest median annual per-patient costs were morbidity-related cost ($17,408), homeless shelter services ($8,618), productivity loss ($8,091), unspecified hospitalization ($6,465), and skilled nursing facilities ($5,143). Ranked by mean cost, the top five were mortality-related cost ($99,532), productivity loss ($17,226), morbidity-related cost ($16,260), skilled nursing facilities ($14,479), and homeless shelter services ($12,977).

## Discussion

This umbrella review represents the most exhaustive overview to date of the economic burden of schizophrenia, drawing on 26 systematic reviews and 152 primary studies. The number of included reviews and primary studies underscores the breadth of primary cost-of-illness literature on schizophrenia. The slight overlap among reviews (CCA of 3.32%) confirms that our umbrella approach draws on a broad and largely non-redundant evidence base.

Most included reviews were rated as “critically low” quality using the AMSTAR 2 checklist, highlighting widespread methodological limitations. However, AMSTAR 2 may not be fully suited to economic burden reviews. Designed for systematic reviews of clinical intervention studies, it emphasizes criteria such as risk of bias assessment and meta-analytic rigor, which are often not applicable in cost-of-illness research due to heterogeneous data sources, non-standardized outcomes, and the frequent absence of meta-analysis. As a result, low scores may reflect a poor fit between the tool and the review type rather than true methodological flaws. Key items like risk of bias assessment and publication bias were often unmet, not necessarily due to inadequate reporting but due to limited relevance. These findings point to the need for appraisal tools better tailored to the structure and aims of economic evaluations.

Across the forty-three primary studies the median per-patient total costs were $33,236 (IQR $14,737-$52,919; mean $47,872). Direct costs were median $23,126 (70% of the level of total costs), of which the largest share was for medical services (median $19,543), while ancillary supports added a modest $1,152. Indirect costs were similar to direct costs, with a median $21,333, which amounted to 64% of the level of total costs. These findings are consistent with the 50% - 90% range reported in Kotzeva et al.’s ten-country synthesis^53^, the 50% - 85% span in Chong et al.’s global review^32^, and exceeds the 44% European mean calculated by Fasseeh et al.^35^

Country income level was found to be the strongest determinant of costs. High-income countries (n = 36) posted a median annual total of $34,175 (mean $56,353), with direct- and indirect medians of $23,985 and $24,436 respectively, illustrating that in this income group direct and indirect costs are similar in magnitude. Within this group, however, regional heterogeneity was striking: North America centred on $33,692, Europe-and-Central-Asia rose to $47,801, and East-Asia-and-Pacific to $38,361. Sub-regional European medians spanned almost an order of magnitude, from $15,357 in Southern Europe to $111,011 in Northern Europe; the Swedish subset alone ranged from $99,183 to $117,067, while Norway peaked at $365,434. These figures mirror the cost dispersion described by Kovács et al.^38^, whose European compilation ranged from €533 to €13,704 and likewise identified hospitalisation as the principal direct driver. In contrast, upper-middle-income settings registered a median total of $3,345, and lower-middle-income settings $3,452; China, for example, averaged $2,943. Two Nigerian studies produced a median $5,932 and recorded direct ($1,477) and indirect ($1,979) costs, underlining how under-resourced health systems and fragmentary records compress the visible burden. No cost study was located for the Middle East or North Africa.

Examining the composition of spending helps understanding these differences. Unspecified hospitalisations were the largest direct cost item (median $6,465; mean $12,152), followed by long-term care (median $4,981) and general inpatient stays (median $4,398). Pharmacotherapy costs (median $1,479) were found to be significantly lower than the $17,115 to $28,101 range reported by Martin et al.^40^. Ancillary supports such as community care (median $4,994), sheltered housing ($3,949) and skilled nursing ($5,143) highlight the interface with social-care systems, while homeless-shelter stays ($8,618) and legal services (median $120, but mean $12,596) show how costs spill into housing and justice budgets. On the indirect cost side, morbidity-related productivity loss carried the heaviest median cost ($17,408), followed by formal productivity loss ($8,091) and informal care ($3,638); mortality-related loss averaged almost $100,000, but was reported in only eleven studies, indicating wide uncertainty around premature death. Overall, indirect costs were found to be $20,643, almost identical to the mean annual €20,664 recorded for Europe by Fasseeh et al.^35^ .

Limiting the analysis to publications after 2000 hardly shifts the totals. In high-income settings the median remains $34,175, but direct and indirect medians jump to $25,697 and $24,807, signalling that two decades of second-generation antipsychotics, long-acting injectables and community-care reform have not altered the underlying cost structure.

Our findings suggest that schizophrenia imposes a large economic burden that is heavily concentrated in high-income countries yet dramatically under-characterized in lower-resource settings. The dominance of hospitalization and indirect costs points to critical opportunities for investment in early intervention, adherence support - particularly through long-acting injectables - community-based services, and rehabilitation programs designed to pre-empt costly relapses and mitigate productivity losses. To support policy-relevant decision-making, future economic evaluations must adopt standardized cost-of-illness methodologies, preregister protocols, transparently report all cost components, and expand data collection into under-represented regions, including the Middle East, North Africa, and lower-middle-income countries. Only through such concerted, high-rigor efforts can we fully capture the true economic footprint of schizophrenia and guide equitable resource allocation to improve outcomes for individuals and societies worldwide.

## Limitations

The literature on the economic burden of schizophrenia is fragmented, with wide variability in healthcare system organization, care pathways, service availability, and cultural perceptions, all of which influence cost estimates and limit cross-country comparability. The included reviews differed in design, cost components, and reporting methods. The absence of standardized methodologies and cost definitions hampers the synthesis and interpretation of findings, underscoring the need for consensus on economic evaluation frameworks in mental health.

Most available data originated from high-income countries, with minimal representation from low- and middle-income settings and none from the Middle East and North Africa. This limits the generalizability of our findings and highlights critical geographic gaps in the evidence base. Cost estimates were frequently extracted from observational studies or administrative claims, which may not fully capture the societal and long-term costs of schizophrenia.

While AMSTAR 2 was applied to ensure methodological transparency, this tool was originally designed for systematic reviews of clinical interventions and may not be fully suited to economic burden reviews, potentially leading to under- or overestimated review qualities.

## Conclusions

This umbrella review provides the most comprehensive overview to date of the global economic burden of schizophrenia. Schizophrenia imposes substantial annual costs per patient, driven by hospitalizations, long-term care, pharmacotherapy, and productivity losses, with wide variation by country income level and region. While high-income countries dominate the evidence base and exhibit the highest costs, data from low- and middle-income regions are seldom published.

Direct medical costs account for the majority of expenditures, but indirect costs particularly related to informal caregiving, morbidity, and premature mortality represent a significant of the total burden. Despite two decades of research, the structure of schizophrenia-related costs has remained largely unchanged, underscoring persistent challenges in care delivery, early intervention, and cost containment.

To inform equitable policy and improve economic evaluation in mental health, future studies must adopt standardized costing frameworks, include all relevant cost domains, and extend data collection into underrepresented settings. Strengthening methodological rigor, expanding geographic coverage, and capturing the full societal impact of schizophrenia are critical steps toward guiding sustainable investment and improving outcomes globally.

## Supporting information

Electronic Supplementary Materials

## Authors contributions

All authors contributed to the formulation of the research hypotheses and the development of the research questions. Data processing and analysis were performed by AI, ÁM, and BB. BN and BN supported the development of the manuscript’s concept. AI wrote the original draft of the manuscript, and all other authors reviewed and revised it critically. All authors have read and approved the final version of the manuscript.

## Availability of data and materials

Data is available at the corresponding author upon reasonable request.

## Ethics approval

Not applicable.

## Acknowledgments

The authors would like to acknowledge reviewers who are not individually named as co-authors: Dóra Csötönyi, Ian Mungai Mngolia and Dominik Turcsán.

