## Supplementary material for "The Global Economic Burden of Schizophrenia: An Umbrella Review of Systematic Reviews and Meta-Analyses": Electronic Supplementary Materials

Supplementary Table 1. PRISMA checklist.

| **Section and Topic** | **Item #** | **Checklist item** | **Location where item is reported** |
| --- | --- | --- | --- |
| **TITLE** | | |  |
| Title | 1 | Identify the report as a systematic review. | Title |
| **ABSTRACT** | | |  |
| Abstract | 2 | See the PRISMA 2020 for Abstracts checklist. | Abstract |
| **INTRODUCTION** | | |  |
| Rationale | 3 | Describe the rationale for the review in the context of existing knowledge. | Introduction |
| Objectives | 4 | Provide an explicit statement of the objective(s) or question(s) the review addresses. | Introduction |
| **METHODS** | | |  |
| Eligibility criteria | 5 | Specify the inclusion and exclusion criteria for the review and how studies were grouped for the syntheses. | Methods |
| Information sources | 6 | Specify all databases, registers, websites, organisations, reference lists and other sources searched or consulted to identify studies. Specify the date when each source was last searched or consulted. | Methods |
| Search strategy | 7 | Present the full search strategies for all databases, registers and websites, including any filters and limits used. | Supplementary Materials |
| Selection process | 8 | Specify the methods used to decide whether a study met the inclusion criteria of the review, including how many reviewers screened each record and each report retrieved, whether they worked independently, and if applicable, details of automation tools used in the process. | Methods |
| Data collection process | 9 | Specify the methods used to collect data from reports, including how many reviewers collected data from each report, whether they worked independently, any processes for obtaining or confirming data from study investigators, and if applicable, details of automation tools used in the process. | Methods |
| Data items | 10a | List and define all outcomes for which data were sought. Specify whether all results that were compatible with each outcome domain in each study were sought (e.g. for all measures, time points, analyses), and if not, the methods used to decide which results to collect. | Methods |
|  | 10b | List and define all other variables for which data were sought (e.g. participant and intervention characteristics, funding sources). Describe any assumptions made about any missing or unclear information. | Methods |
| Study risk of bias assessment | 11 | Specify the methods used to assess risk of bias in the included studies, including details of the tool(s) used, how many reviewers assessed each study and whether they worked independently, and if applicable, details of automation tools used in the process. | Methods |
| Effect measures | 12 | Specify for each outcome the effect measure(s) (e.g. risk ratio, mean difference) used in the synthesis or presentation of results. | Not applicable |
| Synthesis methods | 13a | Describe the processes used to decide which studies were eligible for each synthesis (e.g. tabulating the study intervention characteristics and comparing against the planned groups for each synthesis (item #5)). | Methods |
|  | 13b | Describe any methods required to prepare the data for presentation or synthesis, such as handling of missing summary statistics, or data conversions. | Methods |
|  | 13c | Describe any methods used to tabulate or visually display results of individual studies and syntheses. | Methods |
|  | 13d | Describe any methods used to synthesize results and provide a rationale for the choice(s). If meta-analysis was performed, describe the model(s), method(s) to identify the presence and extent of statistical heterogeneity, and software package(s) used. | Methods |
|  | 13e | Describe any methods used to explore possible causes of heterogeneity among study results (e.g. subgroup analysis, meta-regression). | Methods |
|  | 13f | Describe any sensitivity analyses conducted to assess robustness of the synthesized results. | Not applicable |
| Reporting bias assessment | 14 | Describe any methods used to assess risk of bias due to missing results in a synthesis (arising from reporting biases). | Not applicable |
| Certainty assessment | 15 | Describe any methods used to assess certainty (or confidence) in the body of evidence for an outcome. | - |
| **RESULTS** | | |  |
| Study selection | 16a | Describe the results of the search and selection process, from the number of records identified in the search to the number of studies included in the review, ideally using a flow diagram. | Results |
|  | 16b | Cite studies that might appear to meet the inclusion criteria, but which were excluded, and explain why they were excluded. | Results |
| Study characteristics | 17 | Cite each included study and present its characteristics. | Table 1 |
| Risk of bias in studies | 18 | Present assessments of risk of bias for each included study. | Results |
| Results of individual studies | 19 | For all outcomes, present, for each study: (a) summary statistics for each group (where appropriate) and (b) an effect estimate and its precision (e.g. confidence/credible interval), ideally using structured tables or plots. | Not applicable |
| Results of syntheses | 20a | For each synthesis, briefly summarise the characteristics and risk of bias among contributing studies. | Results, Table 1 |
|  | 20b | Present results of all statistical syntheses conducted. If meta-analysis was done, present for each the summary estimate and its precision (e.g. confidence/credible interval) and measures of statistical heterogeneity. If comparing groups, describe the direction of the effect. | Not applicable |
|  | 20c | Present results of all investigations of possible causes of heterogeneity among study results. | Results |
|  | 20d | Present results of all sensitivity analyses conducted to assess the robustness of the synthesized results. | Results |
| Reporting biases | 21 | Present assessments of risk of bias due to missing results (arising from reporting biases) for each synthesis assessed. | - |
| Certainty of evidence | 22 | Present assessments of certainty (or confidence) in the body of evidence for each outcome assessed. | - |
| **DISCUSSION** | | |  |
| Discussion | 23a | Provide a general interpretation of the results in the context of other evidence. | Discussion |
|  | 23b | Discuss any limitations of the evidence included in the review. | Limitations |
|  | 23c | Discuss any limitations of the review processes used. | Limitations |
|  | 23d | Discuss implications of the results for practice, policy, and future research. | Discussion |
| **OTHER INFORMATION** | | |  |
| Registration and protocol | 24a | Provide registration information for the review, including register name and registration number, or state that the review was not registered. | Methods |
|  | 24b | Indicate where the review protocol can be accessed, or state that a protocol was not prepared. | Methods |
|  | 24c | Describe and explain any amendments to information provided at registration or in the protocol. | Methods |
| Support | 25 | Describe sources of financial or non-financial support for the review, and the role of the funders or sponsors in the review. | Declarations |
| Competing interests | 26 | Declare any competing interests of review authors. | Declarations |
| Availability of data, code and other materials | 27 | Report which of the following are publicly available and where they can be found: template data collection forms; data extracted from included studies; data used for all analyses; analytic code; any other materials used in the review. | Declarations |

Supplementary Table 2. Systematic literature search strategy and number of hits in MEDLINE.

| **Search No.** | **Concepts** | **Search string** | **Date of search** | **Number of hits** |
| --- | --- | --- | --- | --- |
| #1 | Disease | ("schizophrenic"[tiab] OR "bleuler's disease"[tiab] OR "hebephrenia"[tiab] OR "paranoia"[tiab] OR "shared paranoid disorder"[tiab] OR "psychotic affective disorder"[tiab] OR "hebephrenic disorder"[tiab] OR "psychoaffective disorder"[tiab]) OR ("Schizophrenia"[mh]) | 2025-04-09 | 128,186 |
| #2 | Economic Burden | ("work loss"[tiab] OR "work impairment"[tiab] OR "societal cost"[tiab] OR "societal care"[tiab] OR "societal burden"[tiab] OR "social impairment"[tiab] OR "social functioning"[tiab] OR "social cost"[tiab] OR "social care"[tiab] OR "social burden"[tiab] OR "resource utilization"[tiab] OR "resource use"[tiab] OR "resource burden"[tiab] OR "public expenditure"[tiab] OR "productivity loss"[tiab] OR "productivity impairment"[tiab] OR "private expenditure"[tiab] OR "patients cost"[tiab] OR "patient time"[tiab] OR "patient expenditure"[tiab] OR "patient costs"[tiab] OR "patient cost"[tiab] OR "out-of-pocket"[tiab] OR "out of pocket"[tiab] OR "occupational impairment"[tiab] OR "non-healthcare cost"[tiab] OR "miss work"[tiab] OR "lost time"[tiab] OR "lost productivity"[tiab] OR "job loss"[tiab] OR "job impairment"[tiab] OR "intangible cost"[tiab] OR "indirect cost"[tiab] OR "income loss"[tiab] OR "household care"[tiab] OR "household burden"[tiab] OR "home responsibilities"[tiab] OR "home care"[tiab] OR "healthcare resources"[tiab] OR "financial consequences"[tiab] OR "family care"[tiab] OR "family burden"[tiab] OR "familial burden"[tiab] OR "expenditure "[tiab] OR "emergency room visit"[tiab] OR "educational impairment"[tiab] OR "economic consequences"[tiab] OR "earnings "[tiab] OR "direct cost"[tiab] OR "days absent"[tiab] OR "cost-benefit analysis"[tiab] OR "cost study"[tiab] OR "cost of illness"[tiab] OR "cost burden"[tiab] OR "cost assessment"[tiab] OR "co-payment"[tiab] OR "copayment"[tiab] OR "co-pay*"[tiab] OR "carer time"[tiab] OR "carer burden"[tiab] OR "caregiver time"[tiab] OR "caregiver cost"[tiab] OR "caregiver burden"[tiab] OR "budget impact"[tiab] OR "absenteeism"[tiab] OR "economic burden"[tiab]) OR ("Economics, Pharmaceutical"[mh] OR "Health Care Costs"[mh] OR "Cost of Illness"[mh] OR "Financial Stress"[mh] OR "Caregiver burden"[mh] OR "Health Expenditures"[mh] OR "Costs and Cost Analysis"[mh] OR "Cost-Benefit Analysis"[mh] OR "Hospitalization"[mh] OR "Length of stay"[mh] OR "Episode of Care"[mh] OR "Sick Leave"[mh]) | 2025-04-09 | 741,770 |
| #3 | Review Type | ((systematic* [tiab] AND review [tiab]) OR Systematic overview* [tiab] OR Cochrane review* [tiab] OR systemic review* [tiab] OR scoping review [tiab] OR scoping literature review [tiab] OR mapping review [tiab] OR Umbrella review* [tiab] OR (review of reviews [tiab] OR overview of reviews [tiab]) OR meta-review [tiab] OR (integrative review [tiab] OR integrated review [tiab] OR integrative overview [tiab] OR meta-synthesis [tiab] OR metasynthesis [tiab] OR quantitative review [tiab] OR quantitative synthesis [tiab] OR research synthesis [tiab] OR meta-ethnography [tiab]) OR Systematic literature search [tiab] OR Systematic literature research [tiab] OR meta-analyses [tiab] OR metaanalyses [tiab] OR metaanalysis [tiab] OR meta-analysis [tiab] OR meta-analytic review [tiab] OR meta-analytical review [tiab] OR meta-analysis [pt] OR ((search* [tiab] OR medline [tiab] OR pubmed [tiab] OR embase [tiab] OR Cochrane [tiab] OR scopus [tiab] or web of science [tiab] OR sources of information [tiab] OR data sources [tiab] OR following databases [tiab]) AND (study selection [tiab] OR selection criteria [tiab] OR eligibility criteria [tiab] OR inclusion criteria [tiab] OR exclusion criteria [tiab]) ) OR systematic review [pt]) | 2025-04-09 | 656,669 |
| #4 | Combined | #1 AND #2 AND #3 | 2025-04-09 | 330 |
| **#5** | **Limit to English** | | **2025-04-09** | **311 (94.24%)** |

Supplementary Table 3. Systematic literature search strategy and number of hits in EMBASE.

| **Search No.** | **Concepts** | **Search string** | **Date of search** | **Number of hits** |
| --- | --- | --- | --- | --- |
| #1 | Disease | ("schizophrenic":ti,ab,kw OR "bleuler disease":ti,ab,kw OR "hebephrenia":ti,ab,kw OR "paranoia":ti,ab,kw OR "shared paranoid disorder":ti,ab,kw OR "psychotic affective disorder":ti,ab,kw OR "hebephrenic disorder":ti,ab,kw OR "psychoaffective disorder":ti,ab,kw OR "schizophrenia":ti,ab,kw) | 2025-04-09 | 212,328 |
| #2 | Economic Burden | ('work loss':ti,ab,kw OR 'work impairment':ti,ab,kw OR 'societal cost':ti,ab,kw OR 'societal care':ti,ab,kw OR 'societal burden':ti,ab,kw OR 'social impairment':ti,ab,kw OR 'social functioning':ti,ab,kw OR 'social cost':ti,ab,kw OR 'social care':ti,ab,kw OR 'social burden':ti,ab,kw OR 'resource utilization':ti,ab,kw OR 'resource use':ti,ab,kw OR 'resource burden':ti,ab,kw OR 'public expenditure':ti,ab,kw OR 'productivity loss':ti,ab,kw OR 'productivity impairment':ti,ab,kw OR 'private expenditure':ti,ab,kw OR 'patients cost':ti,ab,kw OR 'patient time':ti,ab,kw OR 'patient expenditure':ti,ab,kw OR 'patient costs':ti,ab,kw OR 'patient cost':ti,ab,kw OR 'out-of-pocket':ti,ab,kw OR 'out of pocket':ti,ab,kw OR 'occupational impairment':ti,ab,kw OR 'non-healthcare cost':ti,ab,kw OR 'miss work':ti,ab,kw OR 'lost time':ti,ab,kw OR 'lost productivity':ti,ab,kw OR 'job loss':ti,ab,kw OR 'job impairment':ti,ab,kw OR 'intangible cost':ti,ab,kw OR 'indirect cost':ti,ab,kw OR 'income loss':ti,ab,kw OR 'household care':ti,ab,kw OR 'household burden':ti,ab,kw OR 'home responsibilities':ti,ab,kw OR 'home care':ti,ab,kw OR 'healthcare resources':ti,ab,kw OR 'financial consequences':ti,ab,kw OR 'family care':ti,ab,kw OR 'family burden':ti,ab,kw OR 'familial burden':ti,ab,kw OR 'expenditure':ti,ab,kw OR 'emergency room visit':ti,ab,kw OR 'educational impairment':ti,ab,kw OR 'economic consequences':ti,ab,kw OR 'earnings':ti,ab,kw OR 'direct cost':ti,ab,kw OR 'days absent':ti,ab,kw OR 'cost-benefit analysis':ti,ab,kw OR 'cost study':ti,ab,kw OR 'cost of illness':ti,ab,kw OR 'cost burden':ti,ab,kw OR 'cost assessment':ti,ab,kw OR 'co-payment':ti,ab,kw OR 'copayment':ti,ab,kw OR 'co-pay*':ti,ab,kw OR 'carer time':ti,ab,kw OR 'carer burden':ti,ab,kw OR 'caregiver time':ti,ab,kw OR 'caregiver cost':ti,ab,kw OR 'caregiver burden':ti,ab,kw OR 'budget impact':ti,ab,kw OR 'absenteeism':ti,ab,kw OR 'economic burden':ti,ab,kw OR 'economics, pharmaceutical':ti,ab,kw OR 'health care costs':ti,ab,kw OR 'cost of illness':ti,ab,kw OR 'financial stress':ti,ab,kw OR 'caregiver burden':ti,ab,kw OR 'health expenditures':ti,ab,kw OR 'costs and cost analysis':ti,ab,kw OR 'cost-benefit analysis':ti,ab,kw OR 'hospitalization':ti,ab,kw OR 'length of stay':ti,ab,kw OR 'episode of care':ti,ab,kw OR 'sick leave':ti,ab,kw) | 2025-04-09 | 791,790 |
| #3 | Review Types | ((systematic*:ti,ab AND review:ti,ab) OR 'Systematic overview*':ti,ab OR 'Cochrane review*':ti,ab OR 'systemic review*':ti,ab OR 'scoping review':ti,ab OR 'scoping literature review':ti,ab OR 'mapping review':ti,ab OR 'Umbrella review*':ti,ab OR ('review of reviews':ti,ab OR 'overview of reviews':ti,ab) OR meta-review:ti,ab OR ('integrative review':ti,ab OR 'integrated review':ti,ab OR 'integrative overview':ti,ab OR meta-synthesis:ti,ab OR metasynthesis:ti,ab OR 'quantitative review':ti,ab OR 'quantitative synthesis':ti,ab OR 'research synthesis':ti,ab OR meta-ethnography:ti,ab) OR 'Systematic literature search':ti,ab OR 'Systematic literature research':ti,ab OR meta-analyses:ti,ab OR metaanalyses:ti,ab OR metaanalysis:ti,ab OR meta-analysis:ti,ab OR 'meta-analytic review':ti,ab OR 'meta-analytical review':ti,ab OR term:it OR ((search*:ti,ab OR medline:ti,ab OR pubmed:ti,ab OR embase:ti,ab OR Cochrane:ti,ab OR scopus:ti,ab OR 'web of science':ti,ab OR 'sources of information':ti,ab OR 'data sources':ti,ab OR 'following databases':ti,ab) AND ('study selection':ti,ab OR 'selection criteria':ti,ab OR 'eligibility criteria':ti,ab OR 'inclusion criteria':ti,ab OR 'exclusion criteria':ti,ab)) OR term:it) | 2025-04-09 | 786,018 |
| #4 | Combined | #1 AND #2 AND #3 | 2025-04-09 | 610 |
| **#5** | **Limit to English** | | **2025-04-09** | **586 (96.07%)** |

Supplementary Table 4. Systematic literature search strategy and number of hits in Cochrane Library.

| **Search No.** | **Concepts** | **Search string** | **Date of search** | **Number of hits** |
| --- | --- | --- | --- | --- |
| **#1** | Disease | schizophrenic OR "bleuler's disease" OR hebephrenia OR paranoia OR "shared paranoid disorder" OR "psychotic affective disorder" OR "hebephrenic disorder" OR "psychoaffective disorder" OR schizophrenia | 2025-04-09 | 536 |
| **#2** | Economic Burden | "work loss" OR "work impairment" OR "societal cost" OR "societal care" OR "societal burden" OR "social impairment" OR "social functioning" OR "social cost" OR "social care" OR "social burden" OR "resource utilization" OR "resource use" OR "resource burden" OR "public expenditure" OR "productivity loss" OR "productivity impairment" OR "private expenditure" OR "patients cost" OR "patient time" OR "patient expenditure" OR "patient costs" OR "patient cost" OR "out-of-pocket" OR "out of pocket" OR "occupational impairment" OR "non-healthcare cost" OR "miss work" OR "miss school" OR "lost time" OR "lost productivity" OR "job loss" OR "job impairment" OR "intangible cost" OR "indirect cost" OR "income loss" OR "household care" OR "household burden" OR "home responsibilities" OR "home care" OR "healthcare resources" OR "financial consequences" OR "family care" OR "family burden" OR "familial burden" OR "expenditure " OR "emergency room visit" OR "educational impairment" OR "economic consequences" OR "earnings " OR "direct cost" OR "days absent" OR "cost-benefit analysis" OR "cost study" OR "cost of illness" OR "cost burden" OR "cost assessment" OR "co-payment" OR "copayment" OR "co-pay" OR "carer time" OR "carer burden" OR "caregiver time" OR "caregiver cost" OR "caregiver burden" OR "budget impact" OR "absenteeism" OR "economic burden" OR "Health Care Costs" OR "Cost of Illness" OR "Financial Stress" OR "Caregiver burden" OR "Health Expenditures" OR "Costs and Cost Analysis" OR "Cost-Benefit Analysis" OR "Hospitalization" OR "Length of stay" OR "Episode of Care" OR "Sick Leave" OR "Economics, Pharmaceutical" | 2025-04-09 | 8,630 |
| **#3** | **Combined search** | **#1 AND #2**  **Filters:**   - **Title-Abstract-Keyword** - **Word variations have been searched** - **Only the Cochrane Database of Systematic Reviews were searched** | **2025-04-09** | **520** |

Supplementary Table 5. Systematic literature search strategy and number of hits in APA PsycInfo (via EBSCOhost).

| **Search No.** | **Concepts** | **Search string** | **Date of search** | **Number of hits** |
| --- | --- | --- | --- | --- |
| #1 | Disease | TI (schizophrenic OR "bleuler's disease" OR hebephrenia OR paranoia OR "shared paranoid disorder" OR "psychotic affective disorder" OR "hebephrenic disorder" OR "psychoaffective disorder") OR AB (schizophrenic OR "bleuler's disease" OR hebephrenia OR paranoia OR "shared paranoid disorder" OR "psychotic affective disorder" OR "hebephrenic disorder" OR "psychoaffective disorder") OR KW (schizophrenic OR "bleuler's disease" OR hebephrenia OR paranoia OR "shared paranoid disorder" OR "psychotic affective disorder" OR "hebephrenic disorder" OR "psychoaffective disorder") OR (SU Schizophrenia) | 2025-04-09 | 110,763 |
| #2 | Economic Burden | TI("work loss" OR "work impairment" OR "societal cost" OR "societal care" OR "societal burden" OR "social impairment" OR "social functioning" OR "social cost" OR "social care" OR "social burden" OR "resource utilization" OR "resource use" OR "resource burden" OR "public expenditure" OR "productivity loss" OR "productivity impairment" OR "private expenditure" OR "patients cost" OR "patient time" OR "patient expenditure" OR "patient costs" OR "patient cost" OR "out-of-pocket" OR "out of pocket" OR "occupational impairment" OR "non-healthcare cost" OR "miss work" OR "lost time" OR "lost productivity" OR "job loss" OR "job impairment" OR "intangible cost" OR "indirect cost" OR "income loss" OR "household care" OR "household burden" OR "home responsibilities" OR "home care" OR "healthcare resources" OR "financial consequences" OR "family care" OR "family burden" OR "familial burden" OR "expenditure " OR "emergency room visit" OR "educational impairment" OR "economic consequences" OR "earnings " OR "direct cost" OR "days absent" OR "cost-benefit analysis" OR "cost study" OR "cost of illness" OR "cost burden" OR "cost assessment" OR "co-payment" OR "copayment" OR "co-pay*" OR "carer time" OR "carer burden" OR "caregiver time" OR "caregiver cost" OR "caregiver burden" OR "budget impact" OR "absenteeism" OR "economic burden") OR AB("work loss" OR "work impairment" OR "societal cost" OR "societal care" OR "societal burden" OR "social impairment" OR "social functioning" OR "social cost" OR "social care" OR "social burden" OR "resource utilization" OR "resource use" OR "resource burden" OR "public expenditure" OR "productivity loss" OR "productivity impairment" OR "private expenditure" OR "patients cost" OR "patient time" OR "patient expenditure" OR "patient costs" OR "patient cost" OR "out-of-pocket" OR "out of pocket" OR "occupational impairment" OR "non-healthcare cost" OR "miss work" OR "lost time" OR "lost productivity" OR "job loss" OR "job impairment" OR "intangible cost" OR "indirect cost" OR "income loss" OR "household care" OR "household burden" OR "home responsibilities" OR "home care" OR "healthcare resources" OR "financial consequences" OR "family care" OR "family burden" OR "familial burden" OR "expenditure " OR "emergency room visit" OR "educational impairment" OR "economic consequences" OR "earnings " OR "direct cost" OR "days absent" OR "cost-benefit analysis" OR "cost study" OR "cost of illness" OR "cost burden" OR "cost assessment" OR "co-payment" OR "copayment" OR "co-pay*" OR "carer time" OR "carer burden" OR "caregiver time" OR "caregiver cost" OR "caregiver burden" OR "budget impact" OR "absenteeism" OR "economic burden") OR KW("work loss" OR "work impairment" OR "societal cost" OR "societal care" OR "societal burden" OR "social impairment" OR "social functioning" OR "social cost" OR "social care" OR "social burden" OR "resource utilization" OR "resource use" OR "resource burden" OR "public expenditure" OR "productivity loss" OR "productivity impairment" OR "private expenditure" OR "patients cost" OR "patient time" OR "patient expenditure" OR "patient costs" OR "patient cost" OR "out-of-pocket" OR "out of pocket" OR "occupational impairment" OR "non-healthcare cost" OR "miss work" OR "lost time" OR "lost productivity" OR "job loss" OR "job impairment" OR "intangible cost" OR "indirect cost" OR "income loss" OR "household care" OR "household burden" OR "home responsibilities" OR "home care" OR "healthcare resources" OR "financial consequences" OR "family care" OR "family burden" OR "familial burden" OR "expenditure " OR "emergency room visit" OR "educational impairment" OR "economic consequences" OR "earnings " OR "direct cost" OR "days absent" OR "cost-benefit analysis" OR "cost study" OR "cost of illness" OR "cost burden" OR "cost assessment" OR "co-payment" OR "copayment" OR "co-pay*" OR "carer time" OR "carer burden" OR "caregiver time" OR "caregiver cost" OR "caregiver burden" OR "budget impact" OR "absenteeism" OR "economic burden") OR SU("Economics, Pharmaceutical" OR "Health Care Costs" OR "Cost of Illness" OR "Financial Stress" OR "Caregiver burden" OR "Health Expenditures" OR "Costs and Cost Analysis" OR "Cost-Benefit Analysis" OR "Hospitalization" OR "Length of stay" OR "Episode of Care" OR "Sick Leave") | 2025-04-09 | 141,524 |
| #3 | Review Types | (((TI systematic* OR AB systematic*) AND (TI review OR AB review)) OR (TI "Systematic overview*" OR AB "Systematic overview*") OR (TI "Cochrane review*" OR AB "Cochrane review*") OR (TI "systemic review*" OR AB "systemic review*") OR (TI "scoping review" OR AB "scoping review") OR (TI "scoping literature review" OR AB "scoping literature review") OR (TI "mapping review" OR AB "mapping review") OR (TI "Umbrella review*" OR AB "Umbrella review*") OR ((TI "review of reviews" OR AB "review of reviews") OR (TI "overview of reviews" OR AB "overview of reviews")) OR (TI meta-review OR AB meta-review) OR ((TI "integrative review" OR AB "integrative review") OR (TI "integrated review" OR AB "integrated review") OR (TI "integrative overview" OR AB "integrative overview") OR (TI meta-synthesis OR AB meta-synthesis) OR (TI metasynthesis OR AB metasynthesis) OR (TI "quantitative review" OR AB "quantitative review") OR (TI "quantitative synthesis" OR AB "quantitative synthesis") OR (TI "research synthesis" OR AB "research synthesis") OR (TI meta-ethnography OR AB meta-ethnography)) OR (TI "Systematic literature search" OR AB "Systematic literature search") OR (TI "Systematic literature research" OR AB "Systematic literature research") OR (TI meta-analyses OR AB meta-analyses) OR (TI metaanalyses OR AB metaanalyses) OR (TI metaanalysis OR AB metaanalysis) OR (TI meta-analysis OR AB meta-analysis) OR (TI "meta-analytic review" OR AB "meta-analytic review") OR (TI "meta-analytical review" OR AB "meta-analytical review") OR (PT meta-analysis) OR (((TI search* OR AB search*) OR (TI medline OR AB medline) OR (TI pubmed OR AB pubmed) OR (TI embase OR AB embase) OR (TI Cochrane OR AB Cochrane) OR (TI scopus OR AB scopus) OR (TI "web of science" OR AB "web of science") OR (TI "sources of information" OR AB "sources of information") OR (TI "data sources" OR AB "data sources") OR (TI "following databases" OR AB "following databases")) AND ((TI "study selection" OR AB "study selection") OR (TI "selection criteria" OR AB "selection criteria") OR (TI "eligibility criteria" OR AB "eligibility criteria") OR (TI "inclusion criteria" OR AB "inclusion criteria") OR (TI "exclusion criteria" OR AB "exclusion criteria"))) OR (PT "systematic review")) | 2025-04-09 | 128,194 |
| #4 | Combined | #1 AND #2 AND #3 | 2025-04-09 | 250 |
| **#4** | **Limit to English** | | **2025-04-09** | **229 (91.60%)** |

Supplementary Table 6. Quality Assessment of included reviews based on the AMSTAR II checklist.

| Study ID | Q1 | Q2 | Q3 | Q4 | Q5 | Q6 | Q7 | Q8 | Q9 | Q10 | Q11 | Q12 | Q13 | Q14 | Q15 | Q16 | Confidence Rating* |
| --- | --- | --- | --- | --- | --- | --- | --- | --- | --- | --- | --- | --- | --- | --- | --- | --- | --- |
| Adhikari 2024 | Yes | Partial Yes | Yes | Partial Yes | Yes | Yes | No | Partial Yes | Partial Yes | No | No MA | No MA | No | No | No MA | Yes | Critically low |
| Bramante 2023 | Yes | No | No | Partial Yes | Yes | No | No | Partial Yes | No | Yes | No MA | No MA | No | No | No MA | Yes | Critically low |
| Chong 2016 | No | No | No | Partial Yes | Yes | No | No | No | No | No | No MA | No MA | No | Yes | No MA | Yes | Critically low |
| Christensen 2020 | No | Partial Yes | No | Partial Yes | Yes | Yes | No | No | No | No | No MA | No MA | No | No | No MA | Yes | Critically low |
| Correll 2024 | Yes | Partial Yes | Yes | Partial Yes | Yes | Yes | No | Partial Yes | No | No | No MA | No MA | No | Yes | No MA | Yes | Critically low |
| Dilla 2013 | Yes | No | Yes | Partial Yes | No | No | No | Partial Yes | No | No | No MA | No MA | No | Yes | No MA | Yes | Critically low |
| Fasseeh 2018 | No | No | No | Partial Yes | Yes | Yes | No | No | No | Yes | No MA | No MA | No | Yes | No MA | Yes | Critically low |
| Gustavsson 2011 | No | No | Yes | No | No | No | No | No | No | No | No MA | No MA | No | Yes | No MA | Yes | Critically low |
| Jeun 2024 | Yes | Partial Yes | Yes | Partial Yes | Yes | Yes | No | Partial Yes | No | No | No MA | No MA | No | No | No MA | Yes | Critically low |
| Jin 2017 | No | No | No | Partial Yes | Yes | No | No | No | No | No | No MA | No MA | No | Yes | No MA | Yes | Critically low |
| Kappi 2025 | Yes | Partial Yes | No | Partial Yes | Yes | Yes | No | Partial Yes | Partial Yes | No | Yes | No | Yes | No | Yes | Yes | Low |
| Kinoshita 2013 | Yes | Yes | Yes | No | Yes | Yes | Yes | Yes | Yes | Yes | Yes | Yes | Yes | Yes | Yes | Yes | Low |
| Kotzeva 2023 | Yes | No | No | Partial Yes | No | No | No | No | No | No | No MA | No MA | No | Yes | No MA | No | Critically low |
| Kovács 2018 | Yes | No | No | Partial Yes | Yes | No | No | No | No | Yes | No MA | No MA | No | No | No MA | Yes | Critically low |
| Lin 2023 | No | Yes | No | No | Yes | Yes | No | No | No | No | No MA | No MA | Yes | Yes | No MA | Yes | Critically low |
| Marcellusi 2018 | No | Partial Yes | No | Partial Yes | Yes | No | No | No | No | No | No MA | No MA | No | No | No MA | Yes | Critically low |
| Martin 2022 | Yes | No | No | Partial Yes | Yes | No | No | No | No | No | No MA | No MA | No | No | No MA | No | Critically low |
| OkoliCTC 2022 | Yes | No | No | No | No | No | No | Yes | No | No | Yes | No | No | Yes | Yes | Yes | Critically low |
| Ologundudu 2020 | Yes | No | Yes | Partial Yes | No | No | No | Partial Yes | Partial Yes | No | No MA | No MA | Yes | No | No MA | Yes | Critically low |
| Pennington 2017 | No | No | No | Partial Yes | No | No | No | No | No | No | No MA | No MA | No | No | No MA | Yes | Critically low |
| Reilly 2024 | Yes | Yes | No | Yes | Yes | Yes | Yes | Yes | Yes | Yes | Yes | Yes | Yes | Yes | No | Yes | Low |
| Shuler 2014 | No | No | Yes | No | No | No | No | Partial Yes | No | No | No MA | No MA | No | No | No MA | Yes | Critically low |
| Weber 2022 | No | Partial Yes | Yes | Partial Yes | Yes | Yes | No | No | No | Yes | No MA | No MA | No | Yes | No MA | Yes | Critically low |
| Xia 2011 | Yes | Yes | Yes | No | Yes | Yes | Yes | Yes | Yes | Yes | Yes | Yes | Yes | Yes | Yes | Yes | Low |
| Zhang 2018 | Yes | No | Yes | Partial Yes | No | No | No | Partial Yes | Partial Yes | No | No MA | No MA | Yes | Yes | No MA | Yes | Critically low |
| Zhao 2013 | Yes | No | No | Partial Yes | No | No | No | No | No | No | No | No | No | No | No | Yes | Critically low |

* As defined by Shea et al. Box 2

Supplementary Table 7. Total costs, direct costs and indirect costs per country and income group post-2000.

* Income groups and categorized according to the World Bank. Where the cost comes from one study, only the cost is reported.
